# Evaluating knowledge and attitudes towards dengue fever and its vaccination in China: A questionnaire-based study in healthcare practitioners

**DOI:** 10.1101/2025.09.04.25335082

**Authors:** IpMan Lam, Ye Yao, Yuxiang Sun, Yilan Xia, Yihan Lu

**Author notes:** Corresponding authors, IpMan LAM, Pokfulam Road, Hong Kong, China., Yihan LU, 131 Dong’an Road, Shanghai 200032, China. **Author contributions:** IpMan LAM: conceptualization, data curation, formal analysis, funding acquisition, investigation, methodology, resources, software, visualization, writing – original draft preparation, review & editing. Ye YAO: investigation, resources, writing – review & editing. Yuxiang SUN: investigation, resources, writing – review & editing. Yilan XIA: investigation, resources, writing – review & editing. Yihan LU: conceptualization, data curation, investigation, project administration, resources, supervision, writing – review & editing.

## Abstract

Dengue, a mosquito-borne tropical disease caused by the dengue virus, has seen a significant increase in global incidence in recent decades. China has experienced a rapid rise in dengue cases since 2013, posing a serious threat to population health and increased economic burden. This study aimed to evaluate the knowledge and attitudes towards dengue fever and dengue vaccination among healthcare practitioners in China.

A cross-sectional study was conducted using an online questionnaire survey targeting medical professionals in Zhejiang, Yunnan, and Hainan provinces, which are known endemic areas for dengue virus infection in China. The questionnaire assessed participants’ understanding of dengue virus transmission, high-risk groups, symptoms, knowledge of and attitude towards dengue vaccines. Results showed that 98.6% of participants reported having prior knowledge of the dengue virus. The knowledge of at-risk populations was strongly associated with actual dengue knowledge and vaccine attitude, which potentially drives vaccine acceptance. Most respondents (71.0%) held neutral attitudes towards dengue vaccine, followed by 20.8% that held positive attitudes and 8.2% that held negative attitudes. Individuals with neutral attitude were the most critical in determining the vaccine acceptance rate. Among those that accepted vaccine, 56.1% held neutral attitude, and among those that refused vaccine, 84.3% held neutral attitude. Whilst most respondents (87.6%) reported that they would advocate for the vaccination of susceptible populations against dengue, only 7.5% of respondents were aware of the existence of approved dengue vaccines.

This study provides valuable insights into the current state of knowledge and attitudes towards dengue fever and vaccination among healthcare practitioners in China. Association of various knowledge domains with vaccine attitudes and vaccine acceptance shed light on the more impactful direction for emphasis through educational campaigns and vaccine promotion.

**Author summary:** The evaluation on the knowledge and attitudes towards dengue fever and dengue vaccination among healthcare practitioners in China is crucial in determining the future environment on vaccination policy development. As China has not introduced any dengue vaccines, endemic regions rely greatly on non-pharmaceutical interventions and post-infection treatments. This article reveals the misconceptions and concerns of dengue and dengue vaccines in the medical community in China, which are potential and effect targets of targeted interventions and communication directions that public health authorities can work on to ultimately reduce health burden. This study also identifies false knowledge that are commonly shared, providing an effective reference for endemic regions to learn better of the dengue virus as well as the benefits and health requirements of the dengue vaccine before they make the decision for vaccination. We believe that the introduction and research for dengue virus vaccine would bring beneficial impacts for the Chinese population. The findings of this study provide a scientific basis for decision-making and offer valuable insights for designing effective dengue related education campaigns that can maximize their impact on vaccine attitudes and acceptance.

## Introduction

Dengue virus, a member of the Flaviviridae family and the genus Flavivirus, is the etiological agent that causes dengue fever. Dengue fever is a mosquito-borne tropical disease that has a significant impact on global health. The virus is primarily transmitted through the bites of infected female mosquitoes, predominantly the species Aedes aegypti and Aedes albopictus, to a susceptible human.^1^ The geographical distribution of these mosquitoes reflects the regions where dengue virus is endemic, including various parts of the subtropics and tropics, with notable prevalence in urban and semi-urban areas.^1^

Dengue virus infection has a wide spectrum of clinical presentations, ranging from asymptomatic or mild illnesses to severe flu-like symptoms, and is characterized by high fever, headache, pain behind the eyes, muscle and joint pains, and skin rashes.^2^ Some infection cases can also progress to severe dengue, formerly known as dengue hemorrhagic fever (DHF) or dengue shock syndrome (DSS). These severe forms may be life-threatening and are characterized by plasma leakage, fluid accumulation, respiratory distress, severe bleeding, or organ impairment.^2^

The global incidence of dengue has increased dramatically in recent decades. Since January 2024, there were more than two million dengue infection cases and 500 related deaths globally, and an increased 249% of suspected cases in the first two months of 2024 in comparison to the same period in 2023.^3^ The reasons for this incidence rise are multifaceted, including increased urbanization, population growth, increased global tourism after the COVID-19 pandemic, insufficient public health measures on raising awareness and improving water sanitation , and climate change which has resulted in the expansion of the habitable environment for the mosquito vectors. The lack of a specific and effective antiviral treatment for dengue virus infection accentuates the importance of preventive measures and supportive care. The primary prevention strategy focuses on vector control measures to reduce mosquito population and minimize human-vector contact. Additionally, the continuous development and deployment of dengue vaccine are areas of intensive research, despite the extant challenges in vaccine efficacy and safety especially in antibody-dependent enhancement (ADE).^4^

Understanding the biology of the dengue virus, the pathogenesis of the infection, and the complexities of the immune response to the virus is critical to developing effective treatments and preventive strategies. As the global burden of dengue continues to grow, it is imperative for the international community to strengthen surveillance, improve vector control, invest in vaccine research, and ensure that healthcare systems are prepared to manage both the expected and the more severe presentations of the infection.

Since the outbreak of dengue fever in Guangdong in the 1970s, dengue fever epidemics have occurred every year in China, and the number of cases has increased rapidly after 2013.^5^ From 2016 to 2018, the number of reported cases of dengue fever in China increased from 2050 to 5136, more than doubling.^6,7^ In 2019, before the emergence of the SARS-CoV-2, the number of dengue fever infections reached a historical peak of 22,188 cases.^8^ As the pandemic has gradually been mitigated, there is an expected resurgence of dengue cases in China and in the world that paralleled the increased cross-province travel and expanding mosquito habitat.^9^

In 2019, the number of reported dengue fever cases in China increased significantly, which poses a serious threat to population health and increased economic burden. According to statistics, dengue fever cases caused a total of 46,805,064 yuan in direct health spending in 2019.^10^ In comparison, the cost of prevention and control measures was 6,934,378 yuan, including simple and easily feasible measures such as removing wastewater from potted plants and cleaning streets.^10^

In order to effectively prevent and control the spread of dengue fever, vaccination is an important and effective tool. Currently, two quadrivalent dengue vaccines have been approved for marketing in some countries, namely Dengvaxia® by Sanofi Pasteur and Qdenga® by Takeda Pharmaceuticals. Both vaccines offer some protection against the four serotypes of dengue virus but differ in terms of effectiveness, safety, and suitable populations. The effectiveness of Dengvaxia® is mainly reflected in the protection of patients who have recovered from dengue fever. Its overall efficacy is 82%, with 79% efficacy against the endpoint of hospitalization and 84% efficacy in patients with severe dengue fever.^11^ However, Dengvaxia® may increase the risk of severe dengue fever in people who have never been infected with dengue fever, hence is only recommended for people from 6 to 16 years old and with a history of dengue fever infection by the Centers for Disease Control and Prevention (CDC).^12^

Qdenga® provided data to support its effectiveness and safety, and is recommended for a wider range of people with varied infection history. Qdenga® can be administered to anyone aged over 4 years old, regardless of previous dengue infection history, with an overall efficacy of 80% against all dengue serotypes over 12 months, 90% efficacy against endpoint of hospitalization, and 86% efficacy against dengue hemorrhagic fever patients.^13^ Qdenga® is effective against dengue serotypes 1 and 2, with an efficacy of 78% against hospitalized patients with dengue serotype 1 and 100% against hospitalized patients with dengue serotype 2.^13,14^ Dengvaxia® can provide antibody protection for at least 6 years, while Qdenga® can provide immune protection for at least 4.5 years.^15, 16^ Hence, both vaccines require regular booster shots in epidemic regions.

At present, China has not introduced these vaccine products for use in Chinese populations. This study aims to evaluate the knowledge and attitudes towards dengue fever and dengue vaccination among healthcare practitioners in China. The results of this study will not only provide scientific basis and decision-making reference for the future research, development, or introduction of dengue fever vaccines in China but also provide valuable information and suggestions for formulating effective dengue fever prevention and control strategies and measures in the future.

## Methods

### Study Design and Participants

This cross-sectional study employed a questionnaire survey to assess the knowledge and attitudes towards dengue fever and its vaccination among healthcare practitioners in China, collected in December 2023. The target population included medical professionals working in Zhejiang, Yunnan, and Hainan provinces, which are known endemic areas for dengue virus infection. Participants were recruited from various healthcare settings, including Centers for Disease Control and Prevention (CDC), hospitals (both infectious and non-infectious disease units), and community-level healthcare facilities (e.g., preventive health care departments).

### Questionnaire Development and Administration

The questionnaire was developed by the research team to assess participants’ basic information, understanding of dengue virus and vaccines, and attitudes towards dengue vaccination. The questionnaire consisted of three main sections:

1. Demographics: This section collected data on participants’ nature of work unit, age, education level, specific work department or line, and years of working experience.
2. Understanding of dengue virus and vaccines: This section assessed participants’ knowledge of dengue virus transmission routes, epidemic-prone areas in China, high-risk groups for dengue fever and severe dengue, common symptoms of infection, and understanding of dengue vaccines, including global approval status, applicable groups, safety, effectiveness, main characteristics, and antibody-dependent enhancement (ADE) reaction.
3. Attitudes towards dengue vaccination: This section investigated participants’ anticipated recommendations for dengue vaccination among different groups, awareness of the use of dengue vaccine, and acceptance of the vaccine.

The questionnaire was administered online through the Questionnaire Star platform. The online questionnaire was disseminated to institutions and hospitals, allowing participants to access and complete the survey electronically.

### Data Analysis

Categorical variables were presented as frequencies and percentages, while continuous variables were presented as means and standard deviations. Chi-square tests were used to compare categorical variables, and t-tests were employed for continuous variables in the baseline characteristics assessment.

Actual vaccine knowledge was defined as respondents that answered at least 4 out of the 6 questions on basic dengue vaccine knowledge and reported that they know about antibody-dependent enhancement. Positive attitude towards vaccine was determined for those that expressed positivity on 8 out of the 12 vaccine statements. Negative attitude towards vaccine determined for those that expressed negativity on 8 out of the 12 vaccine statements, and the remaining participants were determined as neutral. To evaluate the associations between perceived and actual knowledge of dengue, as well as the relationships between perceived knowledge, actual knowledge, and vaccine attitudes, odds ratios (ORs) and 95% confidence intervals (CIs) were calculated using logistic regression models. In cases where cell values were 0 in the analysis, the Haldane-Anscombe adjustment was applied by adding 0.5 to each cell.

All statistical analyses were performed using SAS software (version 9.4). A two-tailed p-value of less than 0.05 was considered statistically significant. Figures are constructed using R Studio and GraphPad Prism 10.

### Ethical Considerations

This study was approved by the Institutional Review Board (IRB) of the Fudan University School of Public Health (IRB 00002408 and FWA 00002399) under IRB #2023-10-1085. All respondents accessed the online questionnaire and read the informed consent. They clicked “agree” to the informed consent, which demonstrated they provided informed consent, and then filled out the questionnaire.

## Results

A total of 1,047 healthcare practitioners members were enrolled in the study. A substantial majority of 1,032 participants (98.6%) reported having prior knowledge of the dengue virus, while a minority of 15 participants (1.4%) indicated a lack of such knowledge. The years of professional experience averaged 3.1 (SD = 1.5) for the knowledgeable group and 3.7 (SD = 1.4) for the group without dengue knowledge (p=0.119).

Dengue knowledge did not vary by occupational settings significantly (p=0.066). A large proportion of those with and without reported dengue knowledge were distributed between CDC and the community health service or health centers. Educational attainment varied significantly between the two groups (p=0.012). Most participants in both groups had completed undergraduate education, with 74.3% of the dengue-knowledgeable group and 66.7% of those without dengue knowledge falling into this category. There were 5.5% (n=57/1,032) of participants with a postgraduate degree in the knowledgeable group and 6.7% (n=1/15) of participants with a postgraduate degree in the non-dengue-knowledgeable group.

There were no statistically significant differences between the two groups across the assessed baseline variables except for educational level, suggesting a relatively balanced distribution of participants with respect to dengue knowledge.

### 1. Perceived knowledge and actual knowledge on dengue virus infection

Out of the 1,047 participants who completed the questionnaire, a total of 1,035 respondents (98.9%) affirmed knowledge with the dengue virus. To further assess the actual dengue knowledge of the healthcare practitioners, we conducted an analysis comparing the odds of correct responses between those who perceived themselves as knowledgeable and those who did not. Assessing by the factors with statistically significant odds ratio, healthcare practitioners who perceived themselves as well-versed in dengue were more likely to have accurate knowledge about the disease than their peers who did not consider themselves as knowledgeable (OR = 1.380).

The stratification of dengue-related knowledge into specific domains yielded a diverse understanding of dengue by the two knowledge groups (Table 1). The mean OR obtained for each domain was 1.171 for transmission knowledge, 1.849 for mosquito vector knowledge, 0.929 for high at-risk populations for exposure to infection, 1.342 for high at-risk populations for severe dengue, 1.362 for infection symptoms, and 1.854 for potential complications. Participants reported to have perceived knowledge were in general more likely to have the correct knowledge in dengue knowledge, with misconceptions or misunderstanding in specific parameters like how sexual transmission is not a transmission route for dengue virus (OR = 0.269), or how jaundice is not a common symptom of dengue infection (OR = 0.430).

**Table 1.** Likelihood of actual dengue knowledge among participants with perceived dengue knowledge.

| Variables | Says to know dengue, n (%) |  | Says to not know dengue, n (%) |  | Odds Ratio <sup>a</sup><br>(95% CI) |
| --- | --- | --- | --- | --- | --- |
|  | Actually knows | Actually doesn't know | Actually knows | Actually doesn't know |  |
| Knowledge of dengue transmission |  |  |  |  |  |
| Sexual transmission | 922 (89.3) | 110 (10.7) | 15 (100.0) | 0 (0.0) | 0.269 (1.02, 92.96) |
| Mosquito bite | 1024 (99.2) | 8 (0.8) | 15 (100.0) | 0 (0.0) | 3.888 (1.24, X<sup>b</sup>) |
| Direct contact | 839 (81.3) | 193 (18.7) | 12 (80.0) | 3 (20.0) | 1.215 (1.44, 55.44) |
| Indirect contact | 875 (84.8) | 157 (15.2) | 14 (93.3) | 1 (6.7) | 0.575 (1.11, 22.54) |
| Saliva transmission | 891 (86.3) | 141 (13.7) | 14 (93.3) | 1 (6.7) | 0.652 (1.13, 34.25) |
| Blood transmission | 629 (61.0) | 403 (39.1) | 10 (66.7) | 5 (33.3) | 0.817 (1.34, 10.08) |
| Vertical transmission | 104 (10.1) | 928 (89.9) | 1 (6.7) | 14 (93.3) | 1.088 (1.22, 370.56) |
| Human-mosquito | 322 (31.2) | 710 (68.8) | 5 (33.3) | 10 (66.7) | 0.867 (1.36, 11.61) |
| Knowledge of mosquito vectors |  |  |  |  |  |
| *Anopheles sinensis* | 822 (80.2) | 203 (19.8) | 12 (80.0) | 3 (20.0) | 1.132 (1.41, 42.07) |
| *Anopheles anthropophila* | 952 (92.9) | 73 (7.1) | 13 (86.7) | 2 (13.3) | 2.400 (1.84, 12661.54) |
| *Anopheles anopheles* | 960 (93.7) | 65 (6.3) | 13 (86.7) | 2 (13.3) | 2.716 (1.99, 44975.81) |
| *Culex pipiens* | 932 (90.9) | 93 (9.1) | 14 (93.3) | 1 (6.7) | 1.032 (1.21, 274.89) |
| *Culex tritaenorrhynchus* | 896 (87.4) | 129 (12.6) | 15 (100.0) | 0 (0.0) | 0.223 (1.01, 42.73) |
| *Aedes albopictus* | 916 (88.8) | 109 (10.6) | 12 (80.0) | 3 (20.0) | 2.344 (2.02, 2427.48) |
|  | Actually<br>knows | Actually<br>doesn't know | Actually<br>knows | Actually<br>doesn't know |  |
| *Aedes aegypti* | 819 (79.9) | 206 (20.1) | 9 (60.0) | 6 (40.0) | 2.715 (2.69, 1719.73) |
| *Chironomid* | 980 (95.6) | 45 (4.4) | 14 (93.3) | 1 (6.7) | 2.229 (1.50, 217586.40) |
| **Knowledge of high at-risk population for dengue (exposure)** |  |  |  |  |  |
| Elderly | 406 (39.3) | 626 (60.7) | 8 (53.3) | 7 (46.7) | 0.573 (1.24, 4.67) |
| Infants and young children | 425 (41.2) | 607 (58.8) | 8 (53.3) | 7 (46.7) | 0.618 (1.26, 5.27) |
| Pregnant women | 570 (55.2) | 462 (44.8) | 11 (73.3) | 4 (26.7) | 0.483 (1.17, 4.20) |
| Patients with underlying diseases | 434 (42.1) | 598 (58.0) | 8 (53.3) | 7 (46.7) | 0.641 (1.27, 5.60) |
| Persons with immune deficiency | 359 (34.8) | 673 (65.2) | 4 (26.7) | 11 (73.3) | 1.364 (1.58, 59.56) |
| Re-infected persons with new dengue virus serotype | 559 (54.2) | 473 (45.8) | 9 (60.0) | 6 (40.0) | 0.808 (1.34, 9.11) |
| Smoker | 872 (84.5) | 160 (15.5) | 15 (100.0) | 0 (0.0) | 0.175 (1.01, 19.02) |
| Persons with malnutrition | 678 (65.7) | 354 (34.3) | 12 (80.0) | 3 (20.0) | 0.536 (1.18, 5.84) |
| Outdoor workers | 749 (72.6) | 283 (27.4) | 8 (53.3) | 7 (46.7) | 2.333 (2.38, 537.31) |
| Returning travelers | 564 (54.7) | 468 (45.4) | 6 (40.0) | 9 (60.0) | 1.761 (1.91, 122.96) |
| **Knowledge of high at-risk population for severe dengue (severity)** |  |  |  |  |  |
| Elderly | 806 (78.1) | 226 (21.9) | 9 (60.0) | 6 (40.0) | 2.436 (2.43, 795.65) |
| Infants and young children | 693 (67.2) | 339 (32.9) | 7 (46.7) | 8 (53.3) | 2.315 (2.36, 509.43) |
| Pregnant women | 560 (54.3) | 472 (45.7) | 4 (26.7) | 11 (73.3) | 3.032 (2.75, 8752.13) |
| Patients with underlying diseases | 835 (80.9) | 197 (19.1) | 13 (86.7) | 2 (13.3) | 0.783 (1.22, 21.06) |
|  | Actually<br>knows | Actually<br>doesn't know | Actually<br>knows | Actually<br>doesn't know |  |
| Persons with immune deficiency | 845 (81.9) | 187 (18.1) | 13 (86.7) | 2 (13.3) | 0.835 (1.24, 25.78) |
| Re-infected persons with new dengue virus serotype | 522 (50.6) | 510 (49.4) | 7 (46.7) | 8 (53.3) | 1.160 (1.54, 22.65) |
| Smoker | 901 (87.3) | 131 (12.7) | 15 (100.0) | 0 (0.0) | 0.221 (1.01, 41.19) |
| Persons with malnutrition | 420 (40.7) | 612 (59.3) | 4 (26.7) | 11 (73.3) | 1.754 (1.80, 191.39) |
| Outdoor workers | 739 (71.6) | 293 (28.4) | 13 (86.7) | 2 (13.3) | 0.467 (1.13, 6.12) |
| Returning travelers | 825 (79.9) | 207 (20.1) | 14 (93.3) | 1 (6.7) | 0.412 (1.08, 9.26) |
| **Knowledge of infection symptoms** |  |  |  |  |  |
| Fever | 1018 (98.6) | 14 (1.4) | 15 (100.0) | 0 (0.0) | 2.266 (1.14, X <sup>c</sup>) |
| Nausea/Vomiting | 547 (53.0) | 485 (47.0) | 6 (40.0) | 9 (60.0) | 1.648 (1.83, 90.32) |
| Pain behind eyes | 706 (68.4) | 326 (31.6) | 9 (60.0) | 6 (40.0) | 1.481 (1.72, 57.39) |
| Runny nose | 754 (73.1) | 278 (26.9) | 11 (73.3) | 4 (26.7) | 1.060 (1.42, 24.04) |
| Abdominal pain | 433 (42.0) | 599 (58.0) | 3 (20.0) | 12 (80.0) | 2.583 (2.19, 4932.18) |
| Jaundice | 832 (80.6) | 200 (19.4) | 14 (93.3) | 1 (6.7) | 0.430 (1.08, 10.21) |
| Rash | 765 (74.1) | 267 (25.9) | 12 (80.0) | 3 (20.0) | 0.801 (1.27, 14.05) |
| Bleeding gums | 481 (46.6) | 551 (53.4) | 7 (46.7) | 8 (53.3) | 0.989 (1.44, 14.32) |
| Bone and joint pain | 743 (72.0) | 289 (28.0) | 11 (73.3) | 4 (26.7) | 1.005 (1.40, 20.36) |
| **Knowledge of possible infection complications** |  |  |  |  |  |
| Acute myocarditis/ Acute heart failure | 939 (91.0) | 93 (9.0) | 13 (86.7) | 2 (13.3) | 1.861 (1.61, 1472.41) |
|  | Actually<br>knows | Actually<br>doesn't know | Actually<br>knows | Actually<br>doesn't know |  |
| Pulmonary failure | 511 (49.5) | 521 (50.5) | 5 (33.3) | 10 (66.7) | 1.872 (1.94, 198.66) |
| Liver failure | 715 (69.3) | 317 (30.7) | 10 (66.7) | 5 (33.3) | 1.180 (1.52, 28.22) |
| Acute renal failure | 802 (77.7) | 230 (22.3) | 10 (66.7) | 5 (33.3) | 1.824 (1.90, 175.91) |
| Encephalopathy/ Encephalitis | 664 (64.3) | 368 (35.7) | 7 (46.7) | 8 (53.3) | 2.044 (2.14, 244.89) |
| Acute gastroenteritis | 312 (30.2) | 720 (69.8) | 2 (13.3) | 13 (86.7) | 2.342 (1.83, 8859.50) |

<sup>a</sup> Haldane-Ascombe correction is applied by adding 0.5 to all cells for odds ratio calculation. <sup>b</sup> Value too large. $X^b = 3.67e30$ . $X^c = 1.75e17$ .

Within the domain of dengue transmission knowledge, respondents with perceived knowledge were more likely to correctly identify mosquito bite (OR = 3.888) and vertical transmission (mother-to-fetus) (OR = 1.088) as possible routes of dengue transmission and understand that direct contact is not a possible mechanism (OR = 1.215). Within the domain of mosquito vectors, respondents with perceived knowledge were more likely to correctly identify Aedes agypti (OR = 2.715) and Aedes albopictus (OR = 2.344). Respondents with perceived knowledge were also more likely to identify the most common symptoms of dengue infection including fever (OR = 2.266), nausea/vomiting (OR = 1.648), pain behind eyes (OR = 1.481), and abdominal pain (OR = 2.583). Respondents with perceived knowledge were consistently more likely to have correct understanding across all parameters of the domain of potential complications (1.10 < OR < 2.1).

### 2. Vaccine knowledge

There were 79 (7.5%) out of the 1047 respondents that affirmed the existence of approved dengue vaccines in the world (Figure 1). Among these, 39 participants (49.4%) correctly identified the approved dengue vaccines to be either Dengvaxia®, Qdenga®, or both, while 40 participants (50.6%) demonstrated false knowledge (Figure 1). Regarding approved dengue vaccines in China, 297 (28.4%) respondents were correct that there was none, with 24 (2.3%) respondents falsely believing that there was approved dengue vaccine in China and provided incorrect vaccine names (Figure 1).

**Figure 1.**
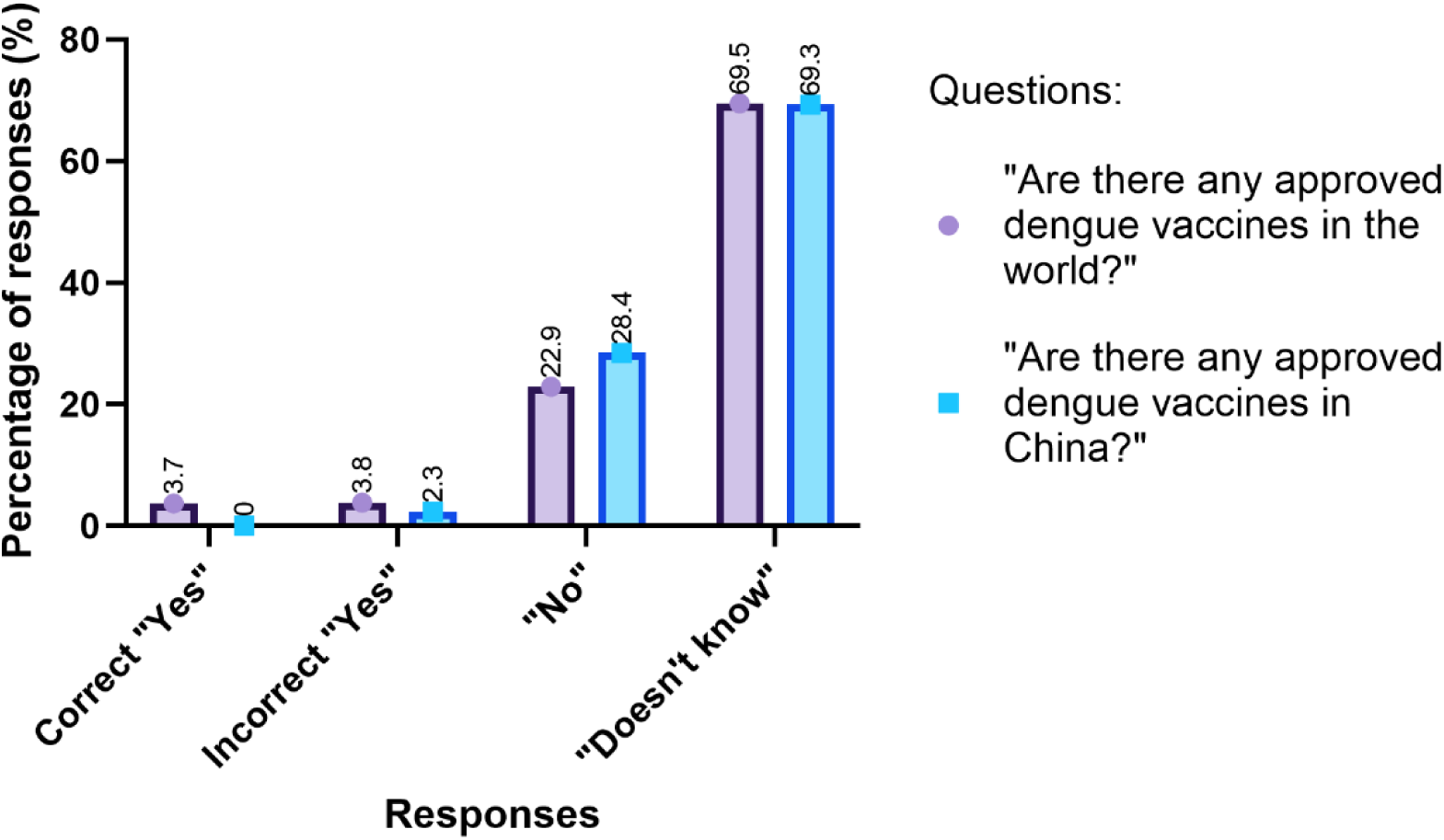
Responses to dengue vaccine approval questions.

Among the 27 respondents who correctly named Dengvaxia as an approved vaccine, 11 respondents selected the correct indication of “9 to 16 years old (recovered)” to be the appropriate age group for vaccination (Figure 2). However, only 4 of the 11 respondents made this selection exclusively (14.8% of the 27 respondents). Among the 11 respondents that correctly named Qdenga as an approved vaccine, 10 respondents selected the correct indication of “older than 4 years old (recovered)” and “older than 4 years old (never infected)” (Figure 2). However, only 2 respondents exclusively selected both choices to reflect that Qdenga can be administered to population aged above 4 years old regardless of dengue infection history (18.1% of the 11 respondents).

**Figure 2.**
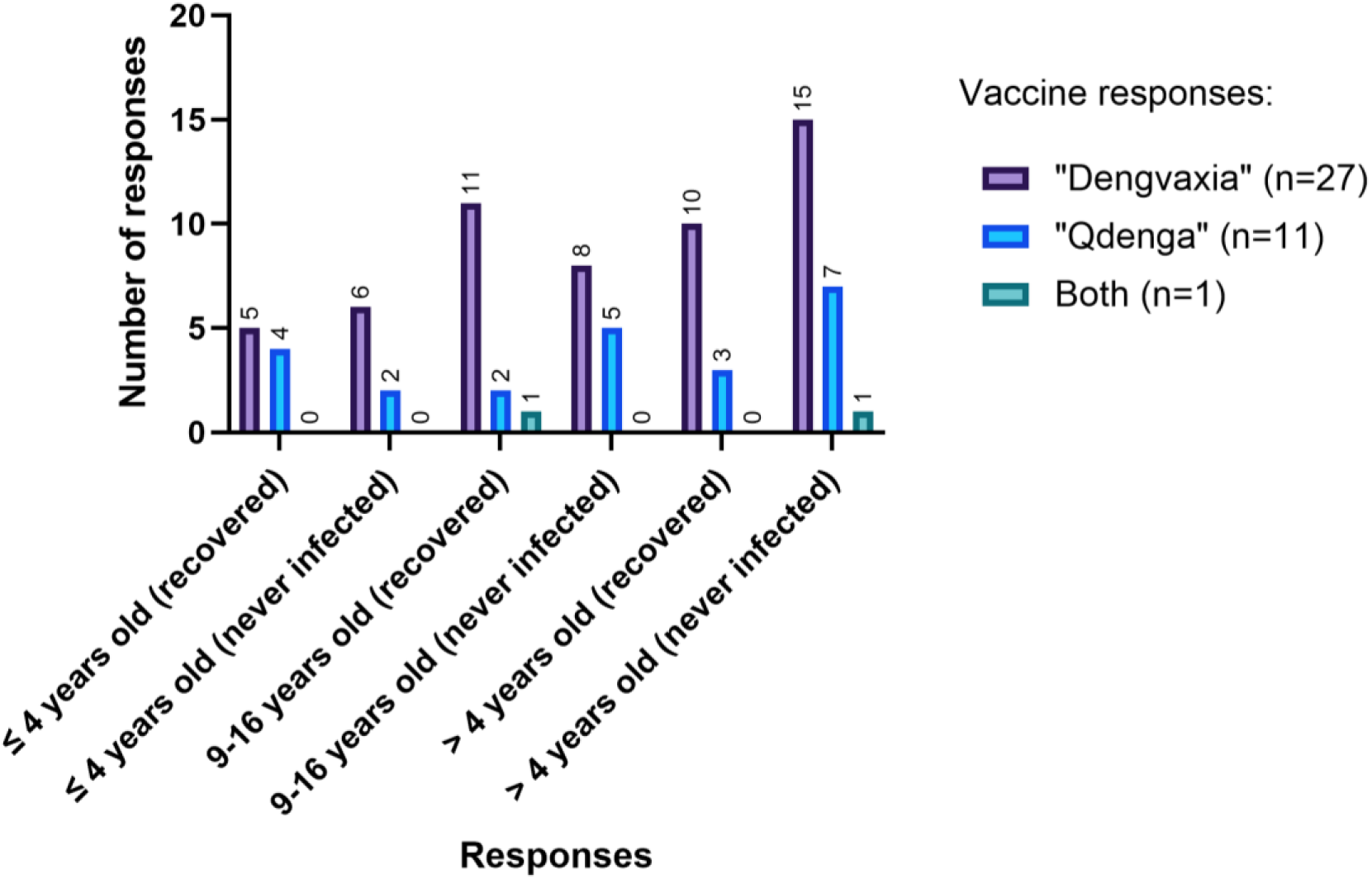
Accurate knowledge on vaccine-appropriate age population.

Only 1 respondent correctly named both vaccines but was not completely accurate on this parameter (Figure 2). However, the selection of “9 to 16 years old (recovered)” and “older than 4 years old (never infected)” reflected good knowledge on vaccine appropriate age population in general.

Majority of the respondents demonstrated good understanding of dengue vaccine’s effect to reduce infection rate, reduce re-infection rate, and reduce hospitalization rate. Agreement proportion was 82.8%, 78.0% and 83.3% respectively (Table 2). However, more than half of the respondents (67.4%) falsely believe that dengue vaccine can prevent dengue virus infection (Table 2). Majority of the respondents were uncertain on the number of doses needed for dengue vaccine administration and whether it offers life-long immunity. 35.1% of the participants correctly understand that dengue vaccine cannot offer life-long immunity against the virus (Table 2).

**Table 2.** Knowledge on dengue vaccine (n=1047)

|  | **"Yes"** | **"No"** | **I don't know** |
| --- | --- | --- | --- |
| **Dengue vaccine can reduce general infection rate** | 867 (82.8) | 25 (2.4) | 155 (14.8) |
| **Dengue vaccine can reduce re-infection rate** | 817 (78.0) | 50 (4.8) | 180 (17.2) |
| **Dengue vaccine can reduce hospitalization rate** | 872 (83.3) | 30 (2.9) | 145 (13.9) |
| **Dengue vaccine can prevent dengue virus infection** | 706 (67.4) | 138 (13.2) | 203 (19.4) |
| **Only 1 dose of dengue vaccine is needed for administration** | 324 (31.0) | 204 (19.5) | 519 (49.6) |
| **Dengue vaccine offers life-long immunity** | 224 (21.4) | 367 (35.1) | 456 (43.6) |

Among the 264 respondents that claimed to know about ADE, 46.2% were correct that not every dengue infection will induce ADE response, 76.9% were correct that ADE response results in severe dengue infection, and 66.7% were correct that ADE response is induced when a recovered patient is re-infected by another serotype of dengue virus (Table 3). 45.8% of the 164 respondents were correct that recovered patients do not develop antibodies against all serotypes, and 56.4% were correct that antibodies in the recovered patients aid the viral invasion of new serotypes of dengue virus into the host (Table 3). In general, the majority of the 264 respondents that claimed to know about ADE demonstrated accurate understanding of ADE associated with dengue virus.

**Table 3.** Knowledge on antibody-dependent enhancement (ADE)

| **Variable** | **Knows antibody-dependent enhancement (ADE)** |  |  |
| --- | --- | --- | --- |
|  | **Actually knows** | **Actually doesn't know** | **Unsure** |
| **Every dengue infection will induce ADE response** | 122 (46.2) | 98 (37.1) | 44 (16.7) |
| **ADE response results in severe dengue infection** | 203 (76.9) | 25 (9.5) | 36 (13.6) |
| **ADE response is induced when a recovered patient is re-infected by another serotype of dengue virus** | 176 (66.7) | 45 (17.1) | 43 (16.3) |
| **Recovered patients develop antibodies against all serotypes** | 121 (45.8) | 110 (41.7) | 33 (12.5) |
| **Antibodies in recovered patients aid the viral invasion of new serotypes of dengue virus into host** | 149 (56.4) | 70 (26.5) | 45 (17.1) |

### 3. Vaccine attitude

Majority of the respondents (87.6%) indicated that they would advocate for the vaccination of susceptible populations against dengue, should a vaccine be approved and accessible in China (Table 4). The inclination to recommend dengue vaccine to the general population and those who have recovered from dengue was less pronounced, with rates of 52.2% and 45.2% respectively (Table 4).

**Table 4.** Dengue vaccine attitudes (n=1047)

| Variables | Very Agree | Agree | No Opinion/<br>I don't know | Disagree | Very Disagree |
| --- | --- | --- | --- | --- | --- |
| **Would you recommend the following to get vaccinated** |  |  |  |  |  |
| More susceptible populations | / | 917 (87.6) | 33 (3.2) | 97 (9.3) | / |
| General population | / | 547 (52.2) | 258 (24.6) | 242 (23.1) | / |
| Dengue-recovered patients | / | 473 (45.2) | 365 (34.9) | 209 (20.0) | / |
| **Attitude towards dengue safety and effectiveness** |  |  |  |  |  |
| Dengue vaccine is harmful to the body | 120 (11.5) | 88 (8.4) | 369 (35.2) | 348 (33.2) | 122 (11.7) |
| Dengue vaccine can effectively reduce infection risk | 324 (31.0) | 460 (43.9) | 224 (21.4) | 31 (3.0) | 8 (0.8) |
| Dengue vaccine has side effects | 81 (7.7) | 250 (23.9) | 554 (52.9) | 130 (12.4) | 32 (3.01) |
| Dengue vaccine will trigger underlying diseases | 73 (7.0) | 94 (9.0) | 492 (47.0) | 312 (29.8) | 76 (7.3) |
| Dengue vaccine has no long-term data to support its safety | 84 (8.0) | 235 (22.5) | 549 (52.4) | 141 (13.5) | 38 (3.6) |
| Dengue vaccine has no long-term data to support effectiveness | 79 (7.6) | 238 (22.7) | 551 (52.6) | 144 (13.8) | 35 (3.3) |
| The human body has its own immunity; dengue vaccination is not necessary | 57 (5.4) | 74 (7.1) | 357 (34.1) | 430 (41.1) | 129 (12.3) |
| Dengue vaccination may serve scientific research purposes | 91 (8.7) | 206 (19.7) | 537 (51.3) | 169 (16.1) | 44 (4.2) |
| Dengue vaccination may serve political purposes | 53 (5.1) | 66 (6.3) | 423 (40.4) | 375 (35.8) | 130 (12.4) |
| If one doesn't go out, one will not be infected with dengue virus; hence dengue vaccine is not needed | 46 (4.4) | 57 (5.4) | 274 (26.2) | 461 (44.0) | 209 (20.0) |
| Dengue vaccination will affect fertility | 49 (4.7) | 48 (4.6) | 450 (43.0) | 367 (35.1) | 133 (12.7) |
| Dengue vaccine may have negative impact on children's development | 50 (4.8) | 57 (5.4) | 486 (46.4) | 331 (31.6) | 123 (11.8) |

Concerning the safety and efficacy of the dengue vaccine, a large percentage (74.9%) of the respondents expressed positivity that the vaccine could diminish the risk of infection. However, concerns regarding adverse effects were noted by 31.6% of participants (Table 4). Further issues included the potential aggravation of underlying diseases (16%), the absence of long-term safety data (30.5%), and a lack of information on long-term efficacy (30.3%) (Table 4). The impact of the vaccine on fertility was a concern for 9.3% of respondents, and a smaller yet considerable fraction of the surveyed population believed that natural immunity provides adequate protection against dengue (12.5%) or expressed the belief that the vaccine is intrinsically harmful (19.9%) (Table 4).

Additionally, some respondents held the view that dengue vaccination campaigns could be driven by motives unrelated to public health, such as research objectives (28.4%) or political agendas (11.4%) (Table 4). Misconceptions were observed, with 9.8% of participants suggesting that the absence of outdoor exposure negates the need for vaccination, and 10.2% expressing concerns about the impact of vaccines on child development (Table 4).

Further analysis was performed to identify the influence of perceived dengue knowledge, actual dengue knowledge, and actual vaccine knowledge on vaccine attitude. Respondents with perceived dengue knowledge were more likely to have a positive attitude towards vaccine, with OR=1.077. However, this association was statistically insignificant, where 95% CI was 0.30 – 3.90.

The association between actual dengue knowledge and vaccine attitude was assessed for all knowledge domains, in which only two of the six domains yielded parameters with statistically significant results (Figure 3). These include knowledge of high at-risk population for severe dengue (severity) that cover immune-deficient individuals and re-infection of new virus serotype, and knowledge of infection symptoms that cover jaundice. Respondents were more likely to have a positive attitude towards dengue vaccine when they knew that persons with immune deficiency were high at-risk of severe dengue infection (OR=1.785, 95% CI: [1.04, 3.05]) compared to those who were not knowledgeable on this. In contrast, respondents who knew re-infected persons who are exposed to new dengue virus serotype were at higher risk of severe dengue infection were less likely to have a positive attitude (OR=0.558, 95% CI: [0.39, 0.81]) compared to those without this correct knowledge. Similarly, respondents that correctly knew jaundice to not be a common symptom were less likely to have a positive attitude (OR= 0.600, 95% CI: [0.37, 0.98]) compared to those without the correct knowledge.

**Figure 3.**
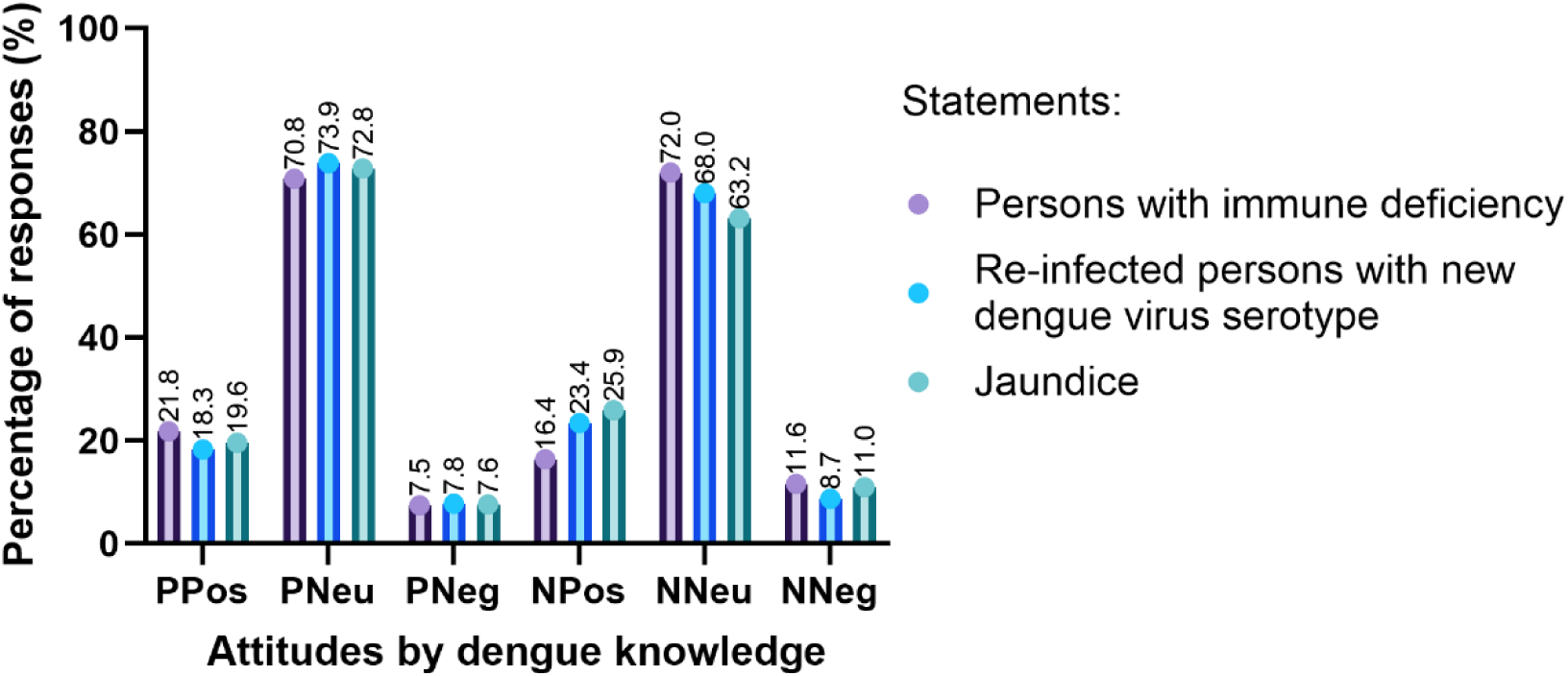
Actual dengue knowledge and vaccine attitude.

PPos = Answered correctly and positive attitude; PNeu = Answered correctly and neutral attitude; PNeg = Answered correctly and negative attitude. NPos = Answered incorrectly and positive attitude; NNeu = Answered incorrectly and neutral attitude; NNeg = Answered incorrectly and negative attitude.

Participant expresses positive attitude towards at least 8 out of 12 parameters. Participant expresses negative attitude towards at least 8 out of 12 parameters. Full table see Supplementary Table 1.

Figure 4 presents the association between actual vaccine knowledge and vaccine attitude. Odds ratio is assessing the likelihood that individuals with vaccine knowledge have a positive attitude compared to a neutral attitude, relative to those without vaccine knowledge. Out of the twelve parameters, only two parameters yielded statistically significant results. Respondents with vaccine knowledge were more likely to have a positive attitude and disagree with the belief that the dengue vaccine is harmful to the body (OR=1.713, 95% CI: [1.06, 2.77]), and believe that the vaccine can effectively reduce infection risk (OR=3.058, 95% CI: [1.52, 6.17]), compared to those without sufficient vaccine knowledge.

**Figure 4.**
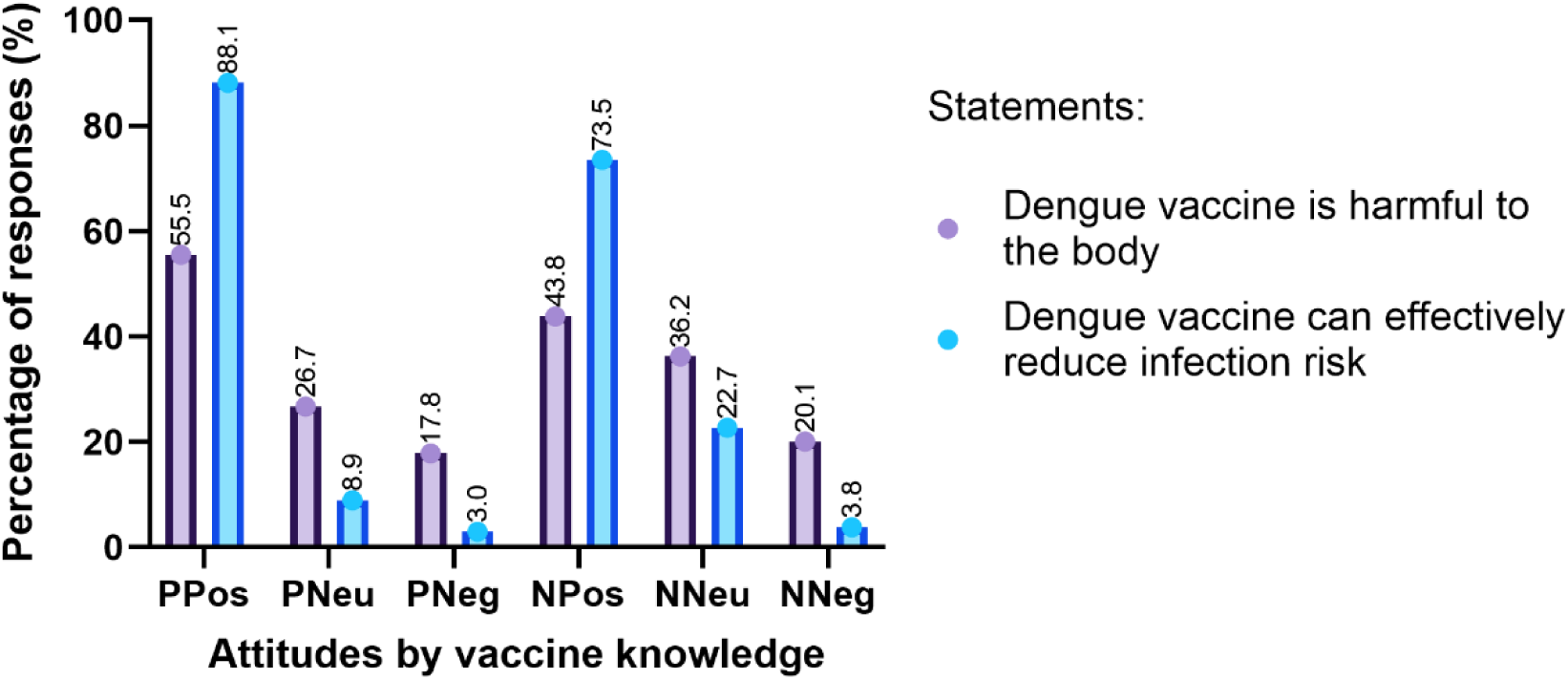
Actual dengue vaccine knowledge and vaccine attitude.

PPos = Knows vaccine and positive attitude; PNeu = Knows vaccine and neutral attitude; PNeg = Knows vaccine and negative attitude. NPos = Does not know vaccine and positive attitude; NNeu = Does not know vaccine and neutral attitude; NNeg = Does not know vaccine and negative attitude.

Know vaccine is defined as those that answered at least 4 out of the 6 questions on basic dengue vaccine knowledge, and says that they know about antibody-dependent enhancement. Full table see Supplementary Table 2.

### 4. Anticipated practice

We assessed the distribution of vaccine acceptance in anticipated practices of vaccination, and the association between vaccine attitude and vaccine acceptance.

If dengue vaccines became available, 48% of respondents would accept vaccination themselves, while 46.8% would recommend it to family and friends. A quarter firmly rejected personal vaccination (25.6%) and recommendations to loved ones (23.4%), with the remainder being unsure (Table 5).

**Table 5.** Anticipated practice on dengue vaccine acceptance (N=1047)

|  | Yes | I don't know | No |
| --- | --- | --- | --- |
| **Will take the dengue vaccine if available** | 503 (48.0) | 276 (26.4) | 268 (25.6) |
| **Will recommend family and friends to take the dengue vaccine if available** | 490 (46.8) | 312 (29.8) | 245 (23.4) |

Through chi-square, we found statistically significance differences (p <0.001) within different levels of vaccine acceptance and within different attitudes towards vaccine. We observed that majority of those that accepted, refused, or had no opinion towards vaccines had a rather neutral attitude towards dengue vaccine (56.1%, 85.1%, 84.3% respectively) (Supplementary Table 3). Respondents that were willing to accept vaccines occupy the highest proportion across all attitudes. 73.9% of those with positive attitude accepted vaccine, 38.0% of those with neutral attitude accepted vaccine, and 69.8% of those with negative attitude accepted vaccine (Table 6). The association between vaccine attitude and vaccine acceptance was confirmed with a yielded OR of 3.946 with 95% CI of 2.623 to 5.937.

**Table 6.** Dengue vaccine attitude by vaccine acceptance (N=1047)

|  | Accept vaccine | No opinion | Refuse vaccine |
| --- | --- | --- | --- |
| **Positive Attitude, n (%)** | 161 (73.9) | 34 (15.6) | 23 (10.6) |
| **Neutral Attitude, n (%)** | 282 (38.0) | 235 (31.6) | 226 (30.4) |
| **Negative Attitude, n (%)** | 60 (69.8) | 7 (8.1) | 19 (22.1) |

## Discussion

This study assessed the perceived and actual knowledge of dengue virus infection and vaccination among healthcare practitioners in China. The results revealed that while a high proportion of participants perceived themselves to be knowledgeable about dengue, their actual knowledge varied across the different domains of dengue knowledge. In general, those who perceived themselves as knowledgeable about dengue were more likely to have accurate knowledge about the disease compared to their peers who did not consider themselves knowledgeable, as indicated by the mean OR of 1.38 and median OR of 1.13.

Looking into the six knowledge domains: transmission mechanisms, mosquito vectors, exposure risk, severe dengue risk, symptoms, and complications yielded mixed results. The general trend demonstrated positive association between perceived and actual knowledge across most domains, including transmission knowledge, mosquito vector knowledge, high at-risk populations for severe dengue, infection symptoms, and potential complications (Table 1). Only one domain had a negative association, where respondents with perceived knowledge were only 0.929 times more likely to have the correct knowledge on the high at-risk populations for exposure to infection. Although statistically significant, the range of the 95% confidence interval was large, indicating a wide range of error may have existed. Looking into the relationship between perceived and actual dengue knowledge in different domains, the most determinant domains of dengue awareness were knowledge on all other groups except for knowledge on high at-risk population to dengue exposure. To have the biggest possible impact, public health initiatives that disseminate information about the dengue virus may find it beneficial to concentrate on these areas or increase education for the populations that are likely to have dengue exposure. This would be beneficial for reducing exposure risks, for example going outdoors without protection.

The investigation also revealed that respondents with perceived dengue knowledge were more likely to have a positive vaccine attitude. Although statistically insignificant, we cannot assume that there was no impact as the results showed a consistent trend, suggesting some underlying pattern between the variables. The trend in the relationship between perceived dengue knowledge and vaccine attitude was potentially promising, affirmed by the positive relationship we yielded for the association between actual dengue knowledge and vaccine attitude (Figure 3, Supplementary Table 1). The general trend reflected a positive association, with only two domains yielding statistically significant parameters which are the most determinant on vaccine attitude (Figure 3, Supplementary Table 1). Respondents with knowledge of persons with immune deficiency being at high risk of severe dengue infection were more likely to have a positive attitude towards the vaccine. This result reflected the findings we obtained when looking into the relationship between perceived and actual dengue knowledge and affirmed knowledge of at-risk populations to be a strong determinant in dengue knowledge and vaccine attitude. Hence, we have obtained a potentially promising direction to focus education campaigns and public awareness promotion projects.

The study findings have important public health implications. If dengue vaccines became available in China, 48% of respondents would accept vaccination, while 46.8% would also recommend it to family and friends. If Qdenga®, with its reported efficacy of 59% against the dengue virus with R_0_ ∼ 1.3, were to be used as the primary vaccine, the current level of acceptance might be considered to have achieved the herd immunity threshold.^17–19^ However, to better prepare for seasonal peaks in dengue cases in endemic provinces as well as considering the risk of severe dengue complications and situation of ADE, it would be advantageous to strive for an even higher acceptance rate. About a quarter firmly rejected personal vaccination (25.6%) and recommendations to loved ones (23.4%). The majority of respondents who accepted, refused, or had no opinion towards vaccines had a rather neutral attitude towards the dengue vaccine (Supplementary Table 3). Specifically, majority of respondents who would refuse the vaccine held a neutral attitude towards the vaccine (84.3%) (Supplementary Table 3). This suggests that vaccine refusal may not always be driven by strong negative attitudes towards the vaccine, but rather by other factors such as misconceptions or personal beliefs. This finding directs a path to focus education campaigns and vaccine promotion. Interestingly, respondents who were willing to accept vaccines occupied the highest proportion across all attitude categories. 73.9% of respondents with positive attitude accepted the vaccine, while 38.0% of those with a neutral attitude and 69.8% of those with a negative attitude accepted the vaccine (Table 6). This finding implies that even among individuals with negative attitudes towards the dengue vaccine, a considerable proportion were still willing to accept vaccination. This could be attributed to various factors, such as the perceived risk of dengue infection, the influence of healthcare providers, or the recognition of the benefits of vaccination despite personal reservations. The study findings also revealed that increasing knowledge about the dengue vaccine was associated to positive attitudes towards its safety and effectiveness, which could ultimately lead to greater vaccine acceptance (Figure 4). Therefore, public health authorities can focus on educating the public and high-risk groups about the risk factors for severe dengue infection and the common symptoms of the disease to promote positive attitudes towards vaccination, and address concerns and misconceptions about the vaccine.

Further studies can be conducted to better understand the complex relationship between knowledge, attitudes, and vaccine acceptance. These studies should aim to identify the key factors influencing vaccine hesitancy and develop evidence-based strategies to overcome barriers to vaccination. From the results in this study, we observed a trend in increased likelihood to have positive attitude towards dengue vaccine when respondents were more knowledgeable in the virus itself, although the association was not statistically significant.

One limitation of the study lies in the unevenly distributed sample size between the two knowledge groups and hence has compromised the power of our findings. Our sample size of participants without perceived dengue knowledge was small (n=15), and participants with knowledge on dengue vaccine was not sufficient (n= 101) when comparing to participants without dengue vaccine knowledge (n=946). This may have affected the statistical power and generalizability of the findings. Future studies should aim to recruit a more balanced and larger sample, which could also potentially resolve the challenge of a wide 95% confidence interval range that we obtained in this study. In terms of validity, the study has higher internal validity but limited external validity because the study was conducted in specific provinces in China and specifically designed for healthcare practitioners, and the findings may not be generalizable to other settings or populations. Future investigations should include a broader range of participants from the dengue endemic regions to assess the consistency of knowledge and vaccine acceptance across different populations.

By increasing awareness and knowledge on determinant knowledge domains, addressing misconceptions and concerns through targeted interventions and effective communication, public health authorities can work towards increasing dengue vaccine uptake and ultimately reducing the burden of dengue disease. We believe that the introduction and research for dengue virus vaccine would bring beneficial impacts for the Chinese population. The findings of this study not only provide a scientific basis for decision-making but also offer valuable insights for designing effective dengue related education campaigns that can maximize their impact on vaccine attitudes and acceptance.

## Data Availability

The raw dataset generated during this study has been deposited in Figshare and will be available upon request once published under the journal, under DOI: [DOI insertion]. Until publication, data access may be provided to peer reviewers or other researchers upon reasonable request by contacting the Ethics Committee at.

## Acknowledgement

We thank all the study respondents who generously completed the online questionnaire. Their contributions have been crucial to the success of this study. I also appreciate my supervisors from Yale, Dr. Inci Yildirim and Prof. Linda Niccolai for their advice and supervision over the final manuscript.

